# BioSignal Copilot: Leveraging the power of LLMs in drafting reports for biomedical signals

**DOI:** 10.1101/2023.06.28.23291916

**Authors:** Chunyu Liu, Yongpei Ma, Kavitha Kothur, Armin Nikpour, Omid Kavehei

## Abstract

Recent advances in Large Language Models (LLMs) have shown great potential in various domains, particularly in processing text-based data. However, their applicability to biomedical time-series signals (e.g. electrograms) remains largely unexplored due to the lack of a signal-to-text (sequence) engine to harness the power of LLMs. The application of biosignals has been growing due to the improvements in the reliability, noise and performance of front-end sensing, and back-end signal processing, despite lowering the number of sensing components (e.g. electrodes) needed for effective and long-term use (e.g. in wearable or implantable devices). One of the most reliable techniques used in clinical settings is producing a technical/clinical report on the quality and features of collected data and using that alongside a set of auxiliary or complementary data (e.g. imaging, blood tests, medical records).

This work addresses the missing puzzle in implementing conversational artificial intelligence (AI), a reliable, technical and clinically relevant signal-to-text (Sig2Txt) engine. While medical foundation models can be expected, reports of Sig2Txt engine in large scale can be utilised in years to come to develop foundational models for a unified purpose. In this work, we propose a system (SignalGPT or BioSignal Copilot) that reduces medical signals to a freestyle or formatted clinical, technical report close to a brief clinical report capturing key features and characterisation of input signal. In its ideal form, this system provides the tool necessary to produce the technical input sequence necessary for LLMs as a step toward using AI in the medical and clinical domains as an assistant to clinicians and patients. To the best of our knowledge, this is the first system for bioSig2Txt generation, and the idea can be used in other domains as well to produce technical reports to harness the power of LLMs. This method also improves the interpretability and tracking (history) of information into and out of the AI models. We did implement this aspect through a buffer in our system.

As a preliminary step, we verify the feasibility of the BioSignal Copilot (SignalGPT) using a clinical ECG dataset to demonstrate the advantages of the proposed system. In this feasibility study, we used prompts and fine-tuning to prevent fluctuations in response. The combination of biosignal processing and natural language processing offers a promising solution that improves the interpretability of the results obtained from AI, which also leverages the rapid growth of LLMs.

## I. INTRODUCTION

The rapid development of modern technology has revolutionized the field of large language models (LLMs). One of the significant advancements in the LLM field in recent years has been the creation of Generative Pre-trained Transformers (GPTs), which have rapidly gained popularity in various fields worldwide [1]. Several theories have been proposed to explain the success of LLMs in diverse applications, some focusing on their architectural advancements and others on using extensive training data. Most prior research has applied LLMs, such as GPT-3, to natural language processing tasks [2]. These studies strongly suggest LLMs’ potential to solve complex domain-specific tasks. While some of the models, such as GPT-4 present multimodal properties, a look at the model repositories and services, such as Hugging Face, reveals the lack of reliable technical BioSignal to Text engineers to be able to increase the reliability of use of these models in medical and clinical domain where preparation of reports per test or procedure is a routine task [3].

Many recent studies have noted the potential of GPT in healthcare. For instance, [4] reports the utilization of ChatGPT in the Medical Knowledge Self-Assessment Program (MKSAP) and its success in helping physicians reduce documentation burdens through the timely provision of the most relevant information. Other studies [5], [6] also mention ChatGPT’s potential use to help generate medical notes for medical consultations and radiological images by the wording of prompts [5], [7]–[3] However, these studies focus on natural language processing or medical images rather, and the application of these models in the medical signal domain remains unattended despite the fact that similar tasks of report preparation and drafting are involved.

The current research on applying natural language processing to biological signal processing is based on the analogy of signals to language and the use of NLP models to help computers learn signal features [34]. Many previous studies have identified the importance of ECG signal feature extraction techniques in identifying cardiovascular diseases [35]–[38] Most previous studies are implemented by combining time-domain, frequency-domain and morphological features with machine learning algorithms [39]. There is a considerable public body of validated clinical and academic knowledge that can be used to harness the power of LLMs and large generative AI models that are growing fast and becoming more domain-specific. This helps make the information flow process in the clinical domain from machines to (expert) humans and back more seamless and interpretable. In the case of ECG, groundbreaking ECG technologies and devices, such as Universal ECG™ [40]–[44], can potentially benefit from obtaining a second-opinion of or an early analysis from an AI machine in particular in case of a junior medical doctor, under pressure emergency department practices, as part of a medical education program or in clinically challenging scenarios.

Ease of use, low cost, wide availability, unprecedented projected growth, and extremely rapid fine-tunability of LLMs for adaptation to particular tasks or domains make integrating them into medical domains as an AI assistant with an expert-in-the-loop necessary [45]. Fig. 1 highlights an overall picture of the rapid growth and expansion of LLMs in terms of parameter size and release date as one of many important factors to highlight the incredible pace at which these languages can be developed, adapted and applied, for instance in legal domain [46]. The data for this graph is extracted from publicly available information. It is also acknowledged that a larger parameter size does not always translate to a better performance [47]. For example, the source and distribution of the training data play a significant role in a successful domain specificity.

**Fig. 1.**
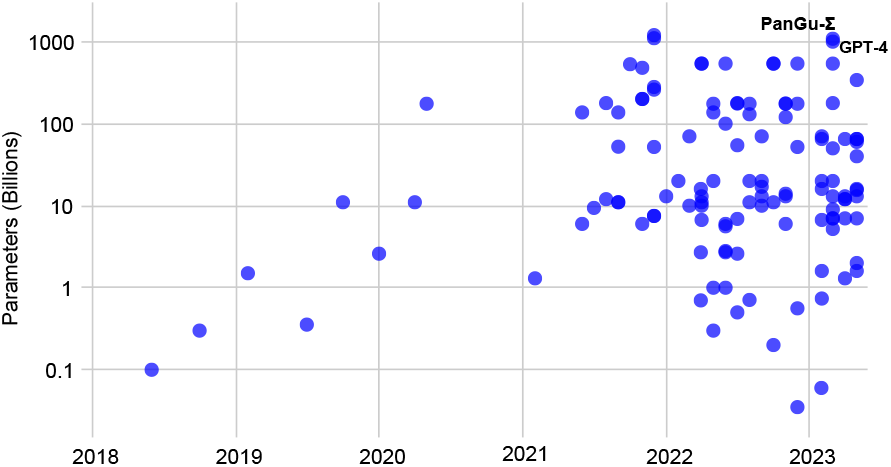
An overall picture of language models release and parameters size, with the publicly accessible GPT-4 [48] and recently reported PanGu-Σ [49] models highlighted.

In this work, we present *SignalGPT*; a system to apply the advance of LLMs to the analysis and assistance in the interpretation of various physiological signals such as electrocardiogram (ECG or EKG), scalp or implanted electroencephalography (EEG), intracranial electrocorticography (ECoG or iEEG), stereo-electroencephalography (sEEG), electrooculography (EOG), electroretinography (ERG), electromyogram (EMG), jugular venous pulse (JVP) monitoring, central venous pressure (CVP) monitoring, and photoplethysmography (PPG) signals. The created system integrates a ChatGPT model with a biomedical signal-processing pipeline.

The engine can process various biomedical data, including ECG, EEG, EMG, etc. It converts the signal or signals into objective and clinical textual descriptions. One of the simplest examples of a feasibility study is ECG, however, it is possible, but more complex to convert an EEG signal into its textual description. It should be noted that the signal-to-text engine is not designed to classify or identify abnormally. The system then inputs the sequence (description) into the fine-tuned ChatGPT model, which evaluates the description alongside the information provided on the subject’s gender, age, comorbidity, and more (e.g. could include height, weight etc). The system then offers its interpretations. Our experiments using ECG signals is only the first step to confirm the viability of this method and through a set of experiments, we were able to demonstrate the feasibility of this approach.

We acknowledge recent works on the imaging domain, either general purpose [3] or specific to medical images [6]. The proposed HuggingGPT [3], is a system that employs a model selector controlled by ChatGPT to solve AI-based tasks. While HuggingGPT comes close to our proposed architecture, we note that HuggingGPT does not extend to biosignals as there is no Hugging Face model for the biosignal-to-text generation to the best of our knowledge. In contrast, SignalGPT is specifically designed to process biomedical signals with a built-in pipeline that specializes in the task. Our model offers a unique advantage over HuggingGPT in this domain. By leveraging the strength of large language models, fine-tuning and combining it with specialized biomedical signal processing pipelines, SignalGPT can provide clinicians with detailed interpretation and analysis of input signals, thereby improving the efficiency and accuracy of clinical decision-making. Furthermore, the interpretability provided by our model enables doctors to understand the reasoning behind SignalGPT’s analysis, promoting greater trust in the system and its output.

## II. METHODS

SignalGPT is a Generative AI-based biomedical signal processing and analyzing system combined with ChatGPT, which is designed for auxiliary clinicians to diagnose the abnormalities of the biomedical signal. SignalGPT consists of Controller (ChatGPT) and the Biomedical Signal Processing Pipeline. The ChatGPT is considered a controller to determine the processing engine in terms of the type of input signal. Another function of the controller is to analyze the description of the given signal based on pre-training on massive corpus and reinforcement learning from human feedback (RLHF). Biomedical Signal Processing Pipeline (BSP) includes a set of signal2text engines for processing different types of medical signals, and these engines integrate corresponding data preprocessing methods and models. The models contain signal-to-text (S2T) generators and signal classifiers. In addition to these two core components, the SignalGPT also provides an interactive interface for users to obtain mandatory information and a gate for fine-tuning ChatGPT by providing feedback.

The workflow of the SignalGPT is demonstrated in Fig. 2. There are five steps.

1. The presented system acquires the gender, age, or other necessary information of the person to be analyzed and diagnosed in the interface from the users. The system in Fig. 2 requires the gender and age of a patient due to ECG as an example. This information can improve the results of ChatGPT interpreting and analyzing signals. At the same time, the path and type of the biomedical signal are required for importing signal data to an appropriate engine.
2. The ChatGPT extracts the type and path of signal from receiving messages and feeds them into the BSP.
3. BSP has a built-in lookup table module to record the number of each engine and the signal types it can handle. The input information finds the corresponding engine through this table and sends the biomedicalsignal-import path into this engine.
4. Signal data are imported following its path, preprocessed, and recognized in the engine generating a prediction of the given biomedical signal and the text which is an objective description of this signal without any opinions. This text is sent back to ChatGPT and analyzed there according to what ChatGPT learned from the corpus. This label is fed into the gate to fine-tune SignalGPT.
5. The generated text is transmitted back to ChatGPT for analysis and interpretation, leveraging its learned knowledge. The ECG signal description is scrutinized by ChatGPT, and its conclusion is extracted to be combined with the prediction produced in step 4 in order to fine-tune ChatGPT at the gate. An eventual reasonable interpretation is exhibited to the user through the interface.

**Fig. 2.**
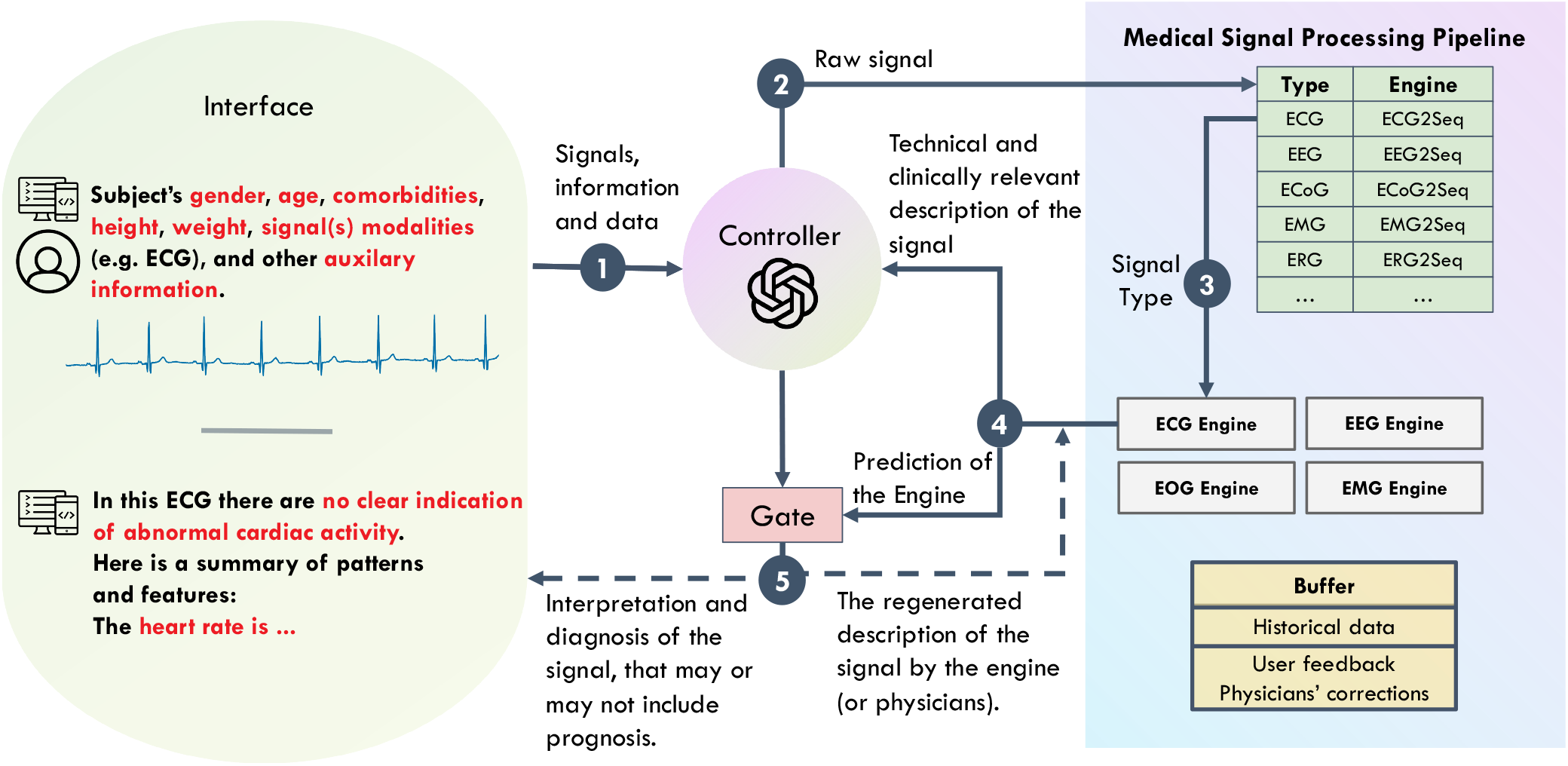
SignalGPT architecture.

In the SignalGPT workflow, we adopt two approaches to fine-tune the output of ChatGPT. (1) We aim to mitigate the impact of the prompt bias in our system, and thus, we standardize the tone of the output generated by the BSP module. In addition, we pose the same question to ChatGPT before sending the produced text to it: “You are a helpful and kind Medical AI Assistant. You can find reasonable explanations online and in your knowledge base based on user descriptions of ECGs and answer the user whether there is any disease. For example, 1st degree AV block (1dAVb), right bundle branch block (RBBB), left bundle branch block (LBBB), sinus bradycardia (SB), atrial fibrillation (AF) and sinus tachycardia (ST).” Not only does the use of consistent language help to avoid misleading ChatGPT but also the posed question reduces the incidence of incorrect responses. (2) After the diagnosis is generated by the ChatGPT, it is matched with the predicted label of the ECG engine of the BSP at the gate. If the diagnosis matches the engine’s label, it is directly outputted to the user interface, and we collect the feedback from users to fine-tune ChatGPT. However, if there is a discrepancy, ChatGPT is used to re-diagnose based on the updated description. This text is regenerated by the engine or fixed by the physician until a reasonable diagnosis is obtained.

We also emphasize the importance of the buffer in SignalGPT. It plays a critical role in optimizing the system to learn and recall patient information like an actual physician. To achieve this objective, we partitioned the cache into two segments. One segment stores the patient’s historical data for a comprehensive review and further analysis by the system. The other segment preserves the user feedback and physician corrections, which are then used by the Engine to enhance its performance via automatic iteration for human feedback reinforcement learning. The physician corrections are used to improve the S2T generator, while the user feedbacks are used to fine-tune the ChatGPT. The frequency of iteration is determined by the size of the buffer. Whenever the buffer is full, the Engine is iterated, and the buffer is cleared in preparation for the next cycle.

The SignalGPT workflow highlights the significance of choosing an appropriate engine. A reliable and high-performing engine can significantly enhance the fine-tuned ChatGPT’s interpretation accuracy and the SignalGPT’s availability. And the physicians can intervene in step 4 to modify the text to increase the accuracy of ChatGPT interpretation.

## III. EXPERIMENTAL SETUP

To validate the feasibility of our system, we conduct experiments utilizing electrocardiogram (ECG) data. To compare the interpretability of ECG analysis using SignalGPT with that just using ChatGPT, we attempted to input unseen ECG data directly into the ChatGPT. However, we discovered that this approach was not feasible as ChatGPT is unable to read raw ECG data and generates answers based on its training data. Instead, we experimented with inputting the description of the ECG signal into ChatGPT to improve accuracy. This approach proved to be effective, as ChatGPT was able to provide more accurate ECG interpretations and diagnoses based on the description of the signal.

Specifically, we utilize an ECG dataset collected by the Telehealth Network of Minas Gerais (TNMG) from 2010 to 2016 and organized by the CODE (Clinical outcomes in digital electrocardiography) group [50]. And we implement data preprocessing, feature extraction, and event processing using the Neurokit2 library. The selection of ECG data and the use of an established library are deliberate choices aimed at ensuring the robustness and generalizability of our system. By conducting these experiments, we are able to assess the performance of our engine in processing ECG data and demonstrate its potential to be applied to other medical signals.

### A. Dataset

The test dataset used in our study comprised 827 12lead ECG records obtained from the Telehealth Network of Minas Gerais (TNMG) in Brazil. The ECG recordings were annotated by two certified cardiologists and a senior specialist [50]. This dataset is publicly available through doi.org/10.5281/zenodo.3625006. This dataset contains 670 normal and 167 abnormal ECG records, of which 1dAVb, RBBB, LBBB, SB, AF, and ST have 28, 34, 30, 16, 13, and 36 records, respectively. The 1dAVb, RBBB, LBBB, SB, AF, and ST are the abbreviations of six types of common ECG abnormalities which is reported in Table I

**TABLE I.**
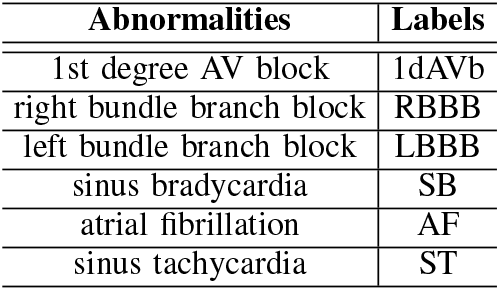
Label specification

| **Abnormalities** | **Labels** |
|---|---|
| 1st degree AV block | 1dAVb |
| right bundle branch block | RBBB |
| left bundle branch block | LBBB |
| sinus bradycardia | SB |
| atrial fibrillation | AF |
| sinus tachycardia | ST |

### B. ECG Engine

Our experimental ECG Engine consists of two models. One is designed for generating the objective text of biomed-ical signals and the other is a classifier to predict the labels of ECG abnormalities.

Firstly, the ECG engine employs Neurokit 0.2.4 to preprocess the raw ECG signal and perform waveform segmentation and feature extraction [51]. Fig. 3 displays the normal ECG signal from a female in her mid-20s on lead V4. The entire ECG record is segmented by heartbeat and features of the ECG waveform, such as the onset and peak of the P waves, the onset and each cusp in the QRS complex, and the onset and peak of the T waves, are identified on each heartbeat. Subsequently, all heartbeats are overlaid and averaged to obtain summarized features for the lead [51].

**Fig. 3.**
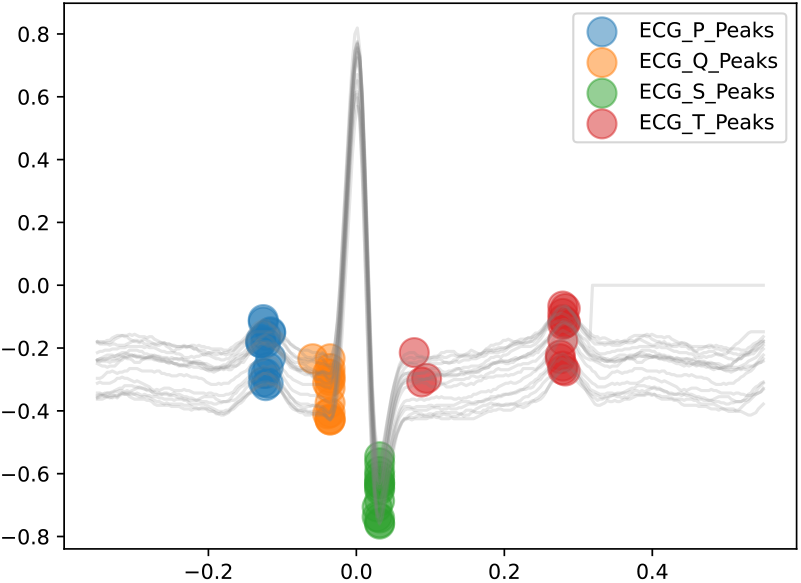
Visualization of waveform segmentation of a normal ECG on lead V4 using Neurokit.

The extracted features are utilized to characterize the input ECG signal in an objective manner. To ensure that the signal’s interpretation and analysis by ChatGPT are unbiased, we have endeavored to maintain a strictly factual and quantitative description. This approach is supported by the inclusion of additional numerical values in the description, which serves to provide a reliable and evidence-based foundation for ChatGPT’s assessment of the ECG signal.

Secondly, the current study utilizes a deep learning network as a classifier which is introduced by [50] to validate the applicability of SignalGPT. The model proposed in [50] employed a classical deep neural network (DNN) to learn features of ECG recordings on their private 12-lead ECG dataset. It is not only capable of recognizing multi-lead features but also has excellent performance. Therefore, we adopted this model as our ECG engine in our proposed system.

### C. Performance Metrics

Given the imbalanced distribution of abnormal and normal samples, this study employs sensitivity (TPR), false positive rate (FPR). The expressions of TPR and FPR are Eq. 1 and Eq. 2 to evaluate the performance of the system in abnormal samples.

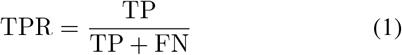

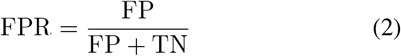

## IV. RESULTS

In this section, we provide two representative examples of ECG diagrams to showcase the system’s functionality. Specifically, the examples include an ECG reading from a healthy female in her mid-20s and another from a male in his mid-70s with atrial fibrillation (AF). We then evaluate SignalGPT’s overall performance on the entire dataset and analyze the obtained results along with any associated system limitations.

### A. Case Study

In this section, we present an analysis of the SignalGPT system through the use of two cases, highlighting its advantages. In the article, we referenced the age range of the subjects due to publication rules, despite having utilized their exact ages in our experiment.

*1) Normal ECG of a female subject in her mid-20s:* The first case is an ECG from a healthy female in her mid-20s, as shown in Fig. 4(a). The ECG exhibits a regular rhythm with an RR interval of approximately three and a half major divisions. The P wave is closely related to the QRS complex, and the PR interval is about three small grids. The QRS complex is not wide or deformed.

**Fig. 4.**
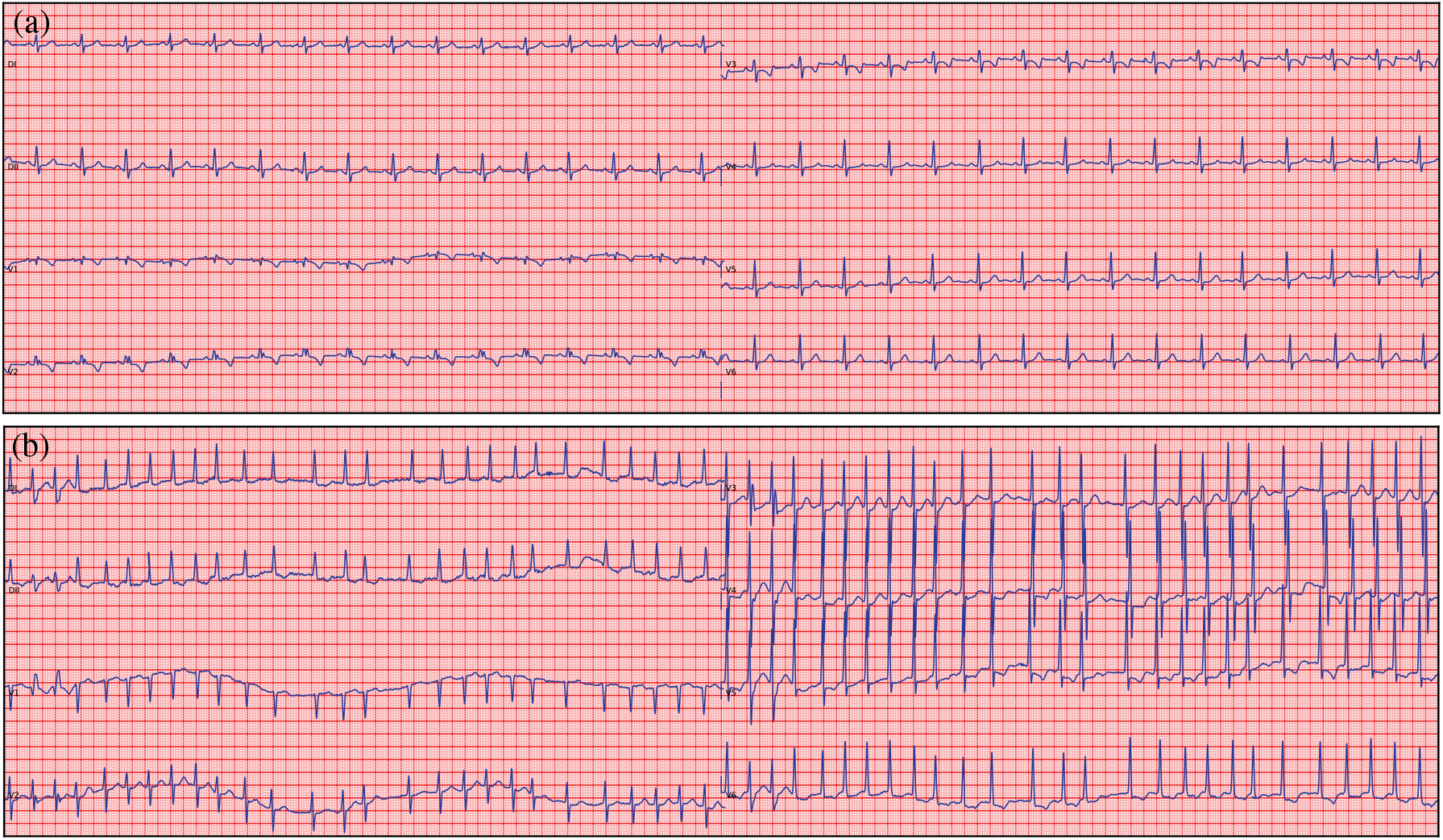
(a) ECG of a female in her mid-20s, which is normal. (b) ECG of a male in his 70s diagnosed as atrial fibrillation (AF). Note that exact ages are masked in this figure and also in the text due to our obligations to disclosure of specific and identifiable information (e.g. age).

#### The description of the output of the ECG engine in SignalGPT is as follows

> *“ This is an ECG of female in her mid-20s. Heart rate is 87. R-R intervals are 0.69 s with a variance of 0.0. P waves last 0.09s with 0.05 mV. PR intervals last 0.12s. The QRS complex lasted 0.09 s with 1.61 mV in limb leads and 1.73 mV in chest leads. In addition, the amplitude of R waves in leads V5 and V6 are 1.86 and 1.7 mV. The maximum of R-wave amplitude in leads I, II and III is 1.3 mV. The other R-wave amplitude in other leads is 1.1 mV. S-wave amplitude in V1 is -0.45 mV. Q-wave amplitude is -0.06 mV. The ST segments last 0.09 s and J-60 points are -0.04 mV. T waves last 0.09s with 0.09 mV. QT intervals last 0.3s.”*

#### To assess the performance of the SignalGPT system in interpreting ECG signals, the output from the ECG engine is entered into ChatGPT. The system interprets the output and provides a response that reads as follows

> *“ Based on the information provided, there are no clear indications of any disease. The ECG shows a normal sinus rhythm with a heart rate of 87 beats per minute. The PR interval, QRS complex, and QT interval are all within normal limits. The R-wave amplitude in leads V5 and V6 are slightly elevated, but this can be a normal variation. The S-wave amplitude in V1 is slightly depressed, but this can also be a normal variation. The ST segments and T waves are also within normal limits. Overall, this ECG appears to be within normal limits and does not suggest any significant cardiac abnormalities or disease. However, it is important to note that a single ECG is not always sufficient to diagnose or rule out cardiac disease, and further testing may be necessary if there are any clinical concerns.”*

#### If ChatGPT reads this unseen ECG data directly, its response is

> *“ The first ECG recording showed sinus tachycardia with a heart rate of about 120 beats per minute. The QRS complex was narrow and regular, sustained. The time is about 80 ms. The PR interval is about 0.16 seconds, which is within the normal range. The QT interval is also normal*.

> *ST-segment elevation in leads V1-V3 suggests acute anterior myocardial infarction. There is also mutual ST-segment depression in leads II, III, and aVF, which may be a sign of inferior myocardial ischemia*.

> *A second ECG recording showed sinus rhythm with a heart rate of approximately 70 beats per minute. The QRS complexes are also narrow and regular, with a duration of approximately 80 milliseconds. The PR interval was about 0.20 seconds, which was within the normal range. The QT interval was slightly prolonged*.

> *T wave inversion in leads V1-V3 may indicate previous anterior myocardial infarction. In addition, ST-segment depression in leads II, III, and aVF suggests inferior wall ischemia*.

> *Taken together, these findings suggest that the patient likely had a history of anterior myocardial infarction and present evidence of inferior wall ischemia on the first ECG recording. A second ECG recording indicated persistent anterior wall ischemia. Further evaluation of additional diagnostic tests, such as cardiac biomarkers and imaging studies, is needed to confirm the diagnosis and guide management.”*

The dataset for each patient includes two types of files to record their invisible ECG data. Therefore, ChatGPT’s answer involves analyzing both the first and second records. We extract the interpretation of SignalGPT and ChatGPT and compare it with the original ECG graph (Fig. 4(a)). Our analysis revealed that SignalGPT provided a more accurate interpretation than the direct use of ChatGPT. Specifically, SignalGPT correctly interprets the ECG as normal, indicating that no significant cardiac abnormalities could be recognized. In contrast, direct ChatGPT’s analysis, while closer to human language, was less accurate in identifying the absence of cardiac abnormalities.

*2) ECG analysis of a male subject in his 70s with a clinical diagnosis of atrial fibrillation:* Moving on to the second case, Fig. 4(b) shows the raw ECG of a male in his 70s. The ECG displays a disappearance of the P wave, which is replaced by the f wave with inconsistent frequency, amplitude, and shape. The equipotential line also disappears, and the RR interval is too short, indicating an irregularly irregular rhythm. Moreover, the amplitude of the QRS complex is high voltage.

The corresponding output of the ECG engine in SignalGPT is shown in the next paragraph. Among them, the inconsistency of the RR interval is expressed by variance. However, the F wave was misdetected as a P wave, despite the apparently unreasonable duration.

> *“ This is an ECG of male in his 70s. Heart rate is 150. R-R intervals are 0.43 s with a variance of 0.011. P waves last 0.04s with 0.03 mV. PR intervals last 0.02s. The QRS complex last 0.08 s with 2.7 mV in limb leads and 5.13 mV in chest leads. In addition, the amplitude of R waves in leads V5 and V6 are 5.13 and 3.62 mV. The maximum of R-wave amplitude in leads I, II and III is 2.29 mV. The other R-wave amplitude in other leads is 3.39 mV. S-wave amplitude in V1 is -1.8 mV. Q-wave amplitude is 0.01 mV. The ST segments last 0.01 s and J-60 points are -0.1 mV. T waves last 0.04s with -0.09 mV. QT intervals disappear.”*

#### The analysis of SignalGPT output to the interface is

> *“ Based on the information provided, this person appears to have atrial fibrillation (AF) with a heart rate of 150 beats per minute. There is also evidence of a prolonged QRS complex duration, with a maximum amplitude of 5.13 mV in chest leads, which may indicate a bundle branch block. However, further evaluation would be necessary to determine whether it is a right bundle branch block (RBBB) or left bundle branch block (LBBB). Additionally, there is evidence of ST segment depression and T wave inversion, which may indicate myocardial ischemia or injury. It is important for this person to seek medical attention for further evaluation and management.”*

#### The interpretation of ChatGPT is shown as follows

> *“This ECG recording shows a sinus rhythm with a rate of approximately 75 beats per minute. The P waves are upright and present before each QRS complex, with a constant PR interval of around 0.20 seconds, suggesting normal atrioventricular conduction. The QRS complexes have a normal duration of around 80 milliseconds*.

> *There are some abnormal findings in this ECG recording. Specifically, there is ST segment elevation in leads II, III, and aVF, which may suggest an acute inferior myocardial infarction. There are also ST segment depressions in leads V1-V3, which may indicate ischemia or injury to the anterior wall of the heart. These findings suggest that the patient may be experiencing an acute coronary syndrome*.

> *There is a normal QT interval, and no signs of atrial or ventricular enlargement or hypertrophy are evident. The axis of the QRS complex is within normal limits, measuring around 60 degrees*.

> *In summary, this hypothetical patient’s ECG shows signs of acute inferior myocardial infarction and possible anterior wall ischemia. Immediate medical attention may be required to manage these conditions. Further diagnostic tests, such as cardiac enzyme testing and echocardiogram, may be needed to confirm the diagnosis and guide treatment.”*

In line with the first case, our proposed SignalGPT was able to detect typical features of atrial fibrillation, such as irregularly irregular rhythms, tachycardia, and irrational P waves. In contrast, the analysis carried out by ChatGPT did not account for these abnormalities. This finding highlights the superiority of SignalGPT in identifying key indicators of atrial fibrillation over ChatGPT.

These two cases further support the superiority of Sig-nalGPT over ChatGPT for biomedical signal analysis. SignalGPT was able to detect important features of the ECG data. However, ChatGPT’s analysis failed to identify abnormalities, highlighting the limitations of directly applying language models to biomedical signal processing. These results demonstrate the need for complementing the biomedical signal processing pipeline to advanced deep learning models like ChatGPT for accurate interpretation and better patient outcomes.

### B. Feasibility verification based on ECG

The results of our study are presented in Table II, which showcases the efficacy of our model in identifying various ECG abnormalities within the test dataset provided in [50]. As illustrated in Table 1, SignalGPT demonstrated perfect sensitivity (100%) for RBBB, LBBB, and SB. The sensitivity for ST was slightly lower at 97.30%. The sensitivities for 1dAVb and AF were the lowest, yet they still surpassed 92%.

**TABLE II.** Performance to Different Types of ECG Abnormalities on TNMG.

| **Abnormalities type** | **TPR (%)** | **FPR (%)** |
|---|---|---|
| **1dAVb** | 92.86 | 1.25 |
| **RBBB** | 100.00 | 0.38 |
| **LBBB** | 100.00 | 0.00 |
| **SB** | 100.00 | 1.48 |
| **AF** | 92.31 | 2.46 |
| **ST** | 97.30 | 1.01 |

Compared to the experimental outcomes obtained by the direct interpretation of ECG signals by ChatGPT, SignalGPT exhibits significantly better performance. The findings of the ChatGPT experiments reveal that the model cannot make precise diagnoses and cannot effectively leverage its knowledge base expertise without prompt engineering support. In contrast, the proposed SignalGPT system demonstrates remarkable performance by utilizing a biomedical signal processing pipeline module that enhances the accuracy of ECG analysis. The experimental outcomes thus illustrate the essential role of an efficient engineering module in facilitating the optimal utilization of the GPT’s inherent knowledge base.

## V. CONCLUSIONS AND DISCUSSIONS

In conclusion, we propose a collaborative system called SignalGPT, which combines multiple biomedical signal processing engines with the state-of-the-art LLM (i.e. ChatGPT). SignalGPT provides a powerful tool for solving task-specific problems in medical signal processing. In particular, the system’s ability to provide detailed interpretation and analysis of input signals can enhance the accuracy and speed of interpretation and annotation, ultimately leading to better clinical and patient outcomes.

One of the critical advantages of SignalGPT is its potential to improve the efficiency of clinicians in making diagnoses and developing treatment plans. With the growing demand for medical services and the increasing complexity of medical data, clinicians are often faced with a significant burden of work. By automating some diagnostic and analytical tasks, SignalGPT can reduce this burden and allow clinicians to focus on other essential aspects of patient care.

Another essential benefit of SignalGPT is its ability to identify patterns and relationships within large amounts of data that might be difficult or impossible to discern using traditional methods. This is particularly important to interpreting medical signals, where slight differences in signal patterns in a particular analysis or relative to historical data can have significant implications for a patient’s situation. SignalGPT can identify these patterns and relationships using advanced machine learning techniques, allowing for more accurate and timely diagnoses.

Overall, the development of SignalGPT represents a significant step forward in medical signal processing. By providing clinicians with a powerful tool for analyzing and interpreting large amounts of data, SignalGPT has the potential to improve the accuracy and speed of preliminary medical interpretation, and ultimately leading to better patient and clinical outcomes. Nonetheless, SignalGPT is not without its limitations. The overall performance of the system is intrinsically tied to the effectiveness of the predictive model. Suboptimal model selection can significantly diminish the accuracy of SignalGPT’s diagnostic reports, potentially influencing the clinical judgments made by physicians. While there is still much work to be done to optimize the system’s performance, the promise of SignalGPT suggests that the future of medical AI is bright.

### A. Human-expert and agent conversational system

As the medical community, industry, academia, and the general public continue to debate the applicability, relevance and reliability of LLMs in the medical field, the power of these emerging tools in “intelligent” report drafting and documentation, as well as their conversational capability can enhance the administrative sides of medical practices, at the very least, and to save time. We demonstrate this feature also in the conceptual application of SignalGPT in EEG reporting in Fig. 5. Building an embedded feature of a *chatbot* into something that was otherwise a patient data management system or conventional AI-assistive technology can create the possibility of Human-expert and agent conversational health care on the way to a possibility for general intelligent machines and/or the creation of more domain-specific foundational models.

**Fig. 5.**
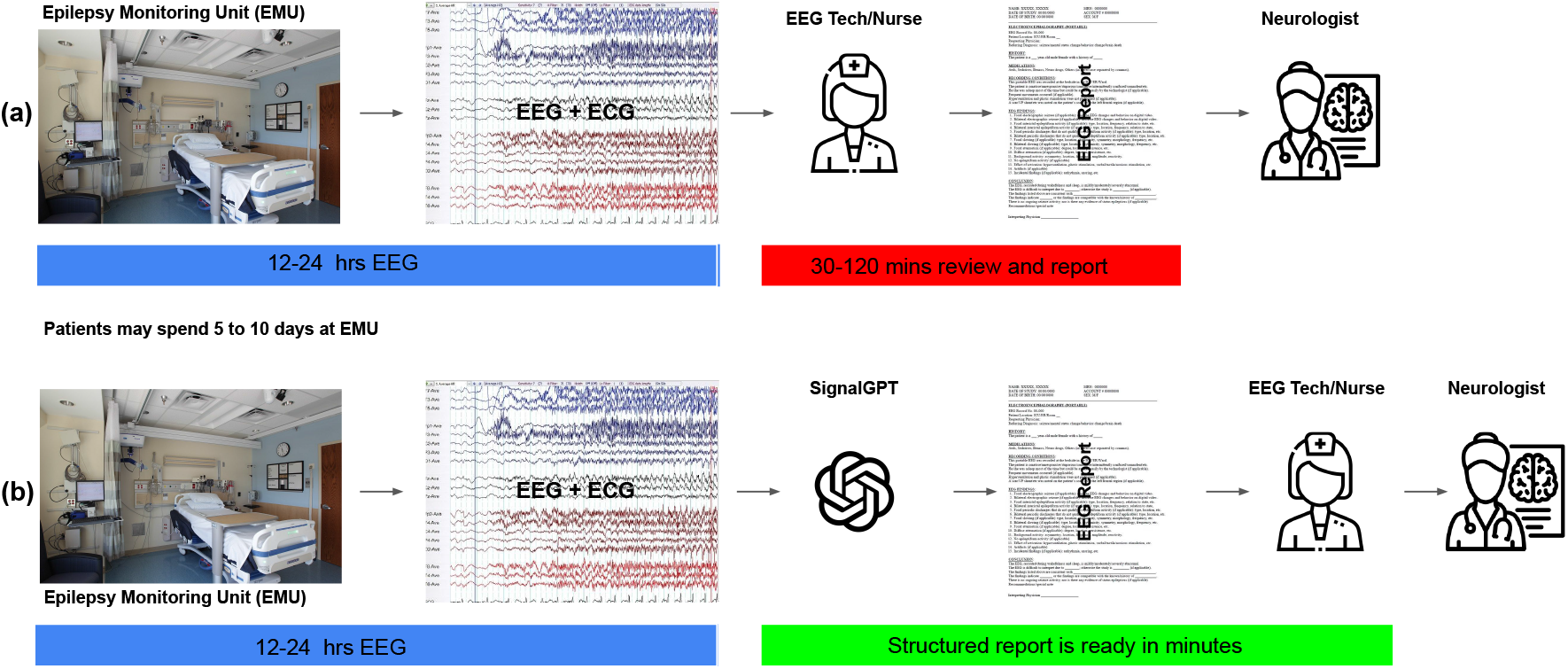
The role of BioSignal Copilot (SignalGPT) in enhancing the electroencephalogram (EEG) report preparation, for instance, in an epilepsy monitoring unit (EMU). (a) Shows EEG and ECG data flow (in sessions, the total can be as long as several days), including evoked potentials studies (Visual Evoked Potential Test, Brain Stem Auditory Evoked Response, Somatosensory Evoked Response), from a testing room to the Technologist and/or Nurses for record keeping and report drafting to the neurologist. (b) Shows an alternative pathway where the time can be saved by using SignalGPT as a *copilot* in the middle.

## Data Availability

All data produced are available online at https://doi.org/10.5281/zenodo.3625006

## Notes

### Competing Interest Statement

The authors have declared no competing interest.

### Funding Statement

This study was partially funded by Children's Hospital at Westmead.

### Author Declarations

The study used (or will use) ONLY openly available human data that were originally located at: https://doi.org/10.5281/zenodo.3625006

